# LENZILUMAB OUTCOMES ACCORDING TO RACE OF COVID-19 PARTICIPANTS IN THE LIVE-AIR PHASE 3 TRIAL

**DOI:** 10.1101/2022.08.18.22278867

**Authors:** Vincent C. Marconi, Adrian Kilcoyne, Franklin Cerasoli, Christopher Polk, Meghan Lewis, Charles D. Burger, Edward Jordan, Cameron Durrant, Dale Chappell, Zelalem Temesgen

## Abstract

**RATIONALE:** The hyperinflammatory immune response of COVID-19, in part orchestrated by granulocyte-macrophage colony-stimulating factor (GM-CSF) can lead to respiratory failure and death with disparities in outcomes between racial subgroups. In the LIVE-AIR trial, the GM-CSF neutralizing antibody lenzilumab improved survival without mechanical ventilation (SWOV) in COVID-19.

**OBJECTIVE:** An analysis of outcomes was performed to determine differences between Black/African American (B/AA) and White participants in LIVE-AIR.

**METHODS:** LIVE-AIR was a phase 3, randomized, double-blind, placebo-controlled trial. Participants hospitalized with COVID-19 pneumonia were randomized 1:1 to receive lenzilumab (1800 mg total) or placebo in addition to standard of care, including remdesivir and/or corticosteroids.

**MEASUREMENTS AND MAIN RESULTS:** Lenzilumab, compared to placebo, numerically improved the likelihood of SWOV (primary endpoint) in B/AA (n=71; 86.8% vs 70.9%; HR, 2.68; 95% confidence interval [CI], 0.88-8.11; p=0.0814) and White (n=343; 85.1% vs 80.8%; HR, 1.41; 95%CI, 0.85-2.35, p=0.182) participants. A statistically significant improvement in SWOV was observed in B/AA (HR: 8.9; 95%CI: 1.08, 73.09; p=0.0418) and White (HR: 2.32; 95%CI: 1.17, 4.61; p=0.0166) participants with baseline CRP<150 mg/L. Lenzilumab numerically, but not statistically, improved secondary endpoints of IMV, ECMO or mortality; ventilator-free days; ICU days and time to recovery in either race while ventilator-free days, ICU days, and time to recovery were statistically improved in B/AA participants with baseline CRP<150 mg/L. Lenzilumab was well tolerated without differences in serious adverse events regardless of race.

**CONCLUSION:** Lenzilumab significantly improved SWOV and some key secondary outcomes in B/AA COVID-19 participants with baseline CRP<150 mg/L. NCT04351152

## INTRODUCTION

COVID-19 results from a hyperinflammatory immune response. Granulocyte-macrophage colony-stimulating factor (GM-CSF) is a key initiator of this response that results in elevated downstream cytokines such as IL-6 and IL-1; the production of inflammatory chemokines such as MCP-1, IL-8, and IL-10 [1]; tissue damaging oxidative stress; and elevated markers of systemic inflammation such as CRP, D-dimer, and ferritin. GM-CSF may therefore be a target for treatment in the early stages of the hyperinflammatory immune response, ultimately preventing the downstream sequelae. Lenzilumab, an anti-human GM-CSF monoclonal antibody that directly binds GM-CSF, with high specificity and affinity, and a slow off rate, prevents GM-CSF signaling through its receptor.[2] The randomized, double-blind, placebo-controlled Phase 3 LIVE-AIR trial demonstrated that lenzilumab improved survival without invasive mechanical ventilation (SWOV) beyond that provided by available treatments, including corticosteroids and/or remdesivir, in patients hospitalized with COVID-19 requiring supplemental oxygen but not yet invasive mechanical ventilation.[3] The response was accentuated in patients with baseline C-reactive protein (CRP) levels less than 150 mg/L, suggesting greater treatment benefit by interrupting the trajectory of less advanced COVID-19 progression.[4]

Racial disparities are apparent in the COVID-19 pandemic. In both the United States and Brazil, the Black population and other racial and ethnic minority groups have experienced significantly higher burden of illness related to COVID-19 compared to the White population, including, more infections, hospitalizations, and a higher mortality rates.[5-7] The disproportionate burden of COVID-19 may result from a more exuberant acute response and exacerbation of chronic inflammation influenced by social determinants of health such as deep-rooted environmental, economic, and political inequalities.[8-10]

The pathophysiology of this hyperinflammatory immune response involving GM-CSF may include but is not limited to disruption of ACE1/ACE2 axis of the renin-angiotensin system, which appears to be more prevalent in B/AA, leading to enhanced inflammation and is postulated to be augmented by SARS-CoV-2 infection[11]; glucose-6-phosphate dehydrogenase (G6PD) deficiency, the prevalence of which is highest among B/AA males[12], resulting in loss of essential oxidant defense during hyperinflammatory conditions[13]; and polymorphism in the Duffy antigen chemokine receptor (DARC) observed in 67% of African Americans[14] leading to cytokine-mediated immune dysregulation[14, 15], which is previously demonstrated to heighten pro-inflammatory status[16] and result in severe acute lung injury.[14, 17]

The purpose of this sub-analysis of LIVE-AIR was to evaluate the efficacy of lenzilumab in both B/AA and White hospitalized COVID-19 patients; particularly in those with baseline CRP levels indicate less advanced COVID-19 progression prior to treatment.

## METHODS

The LIVE-AIR trial design has been previously described in detail.[3] Central IRB (ADVARRA) approval was obtained in addition to local IRB approval at each participating institution as appropriate for all aspects of this company-sponsored phase 3 clinical trial. All necessary patient/participant consent has been obtained and the appropriate institutional forms have been archived. The trial design is briefly summarized here.

### Trial Design

LIVE-AIR was a randomized, double-blind, placebo-controlled, phase 3 trial (NCT04351152) and enrolled hospitalized participants with COVID-19 pneumonia. Eligibility criteria included age 18 years or older, virologically confirmed SARS-CoV-2, and pneumonia diagnosed by chest x-ray or computed tomography. Participants must have been hospitalized with a clinical ordinal score, adapted from the NIH-sponsored Adaptive COVID-19 Treatment Trial (ACTT, NCT 04280705)[18] of 5 (no requirement for oxygen therapy) or clinical ordinal score of 4 (supplemental oxygen in the form of low-flow oxygen) or clinical ordinal score of 3 (high-flow oxygen, or non-invasive positive pressure ventilation) Enrolled participants were randomized 1:1 to receive lenzilumab or matched placebo in addition to current standard treatments per institutional guidelines at each site. Three doses of lenzilumab (total of 1800 mg within a 24-hour period, divided into three equal doses of 600 mg each) or placebo was administered 8 hours apart via a 1-hour IV infusion per dose. Participants were stratified by age (<65 or >65) and disease severity (ordinal score 5 or 4 vs. 3). The primary efficacy endpoint was SWOV by Day 28. For purposes of the survival analysis for the primary endpoint, an event was defined as mortality or the requirement for invasive mechanical ventilation (IMV). Secondary endpoints included time to recovery, the proportion of participants with the composite endpoint of IMV (ordinal score 2), ECMO (ordinal score 2) or death (ordinal score 1); ventilator-free days; duration of ICU; mortality, and safety. For purposes of this sub-analysis only B/AA and White subjects were included.

### Statistical Analysis

The primary endpoint was the difference between lenzilumab treatment and placebo treatment, in addition to standard treatments including remdesivir and steroids, in SWOV through 28 days following randomization in the prespecified modified intent to treat population (mITT) who received at least one dose of investigational treatment under the documented supervision of the principal investigator or sub-investigator. The mITT population was defined as the primary analysis and a Cox proportional hazard model (HR: lenzilumab relative to placebo) accounting for the stratification variables (i.e., age and disease severity) was used, supplemented by a display of K-M curves in each treatment group. The Cox proportional hazard model included the time to first event (death or IMV) as the dependent variable, (1=IMV use or death, 0=alive with no IMV use); treatment (covariate); and strata (covariates). Where data were non-proportional based on a Chi-squared test proposed by Grambsch and Therneau with a global p-value <0.05, a Cox proportional hazard model with weighted extension was used to correct for non-proportionality. Baseline CRP values were determined based on the screening value and if the participant did not have a screening value, then the day 1 value was used.

For each secondary endpoint, the proportion of participants that had the event was calculated by treatment group. An odds ratio was calculated for the composite endpoint of the first incident of IMV, ECMO, or death using logistic regression and including the baseline age group and disease category as covariates. For ventilator-free days and duration of ICU, a nonparametric stratified Wilcoxon test was performed using age strata and disease severity strata as stratification variables. Hazard ratios were calculated for each of time to death and time to recovery, separately, as described above. For time to recovery, deaths were censored at Day 28. Participants who were alive, yet did not recover, were right-censored at the date of the last non-missing assessment of the 8-point clinical status ordinal scale on or prior to Day 28. All data reported herein are reported through Day 28.

## RESULTS

### Participants

Five hundred, twenty-eight participants were screened, of whom 520 were randomized (ITT population) and 479 were included in the pre-specified mITT population (Figure 1). This represented 92% (479/520) of the total population, of which 90% and 94% of each population were randomized to lenzilumab (236/261) and placebo (243/259), respectively. The mITT population only included those who had access to basic supportive care for COVID-19 and received treatment documented by the principal investigator (PI) or sub-investigator.[3] A total of 414 patients were included in this sub-analysis, including 71 who self-identified as B/AA and 343 as White (Figure 1). The remaining excluded participants were Asian (1%), American Indian (3%) or Other (9%) and comprised small percentages of the overall population. Lenzilumab treatment included 165 White and 38 B/AA participants, while placebo treatment included 178 White and 33 B/AA participants. Since differential response to lenzilumab was observed in participants with baseline CRP <150 mg/L and ≥ 150 mg/L[4], the current evaluation stratified participants in those who had documented baseline CRP levels, that excluded 43 participants (18%) in the lenzilumab group and 46 (19%) in the placebo group.

**Figure 1.**
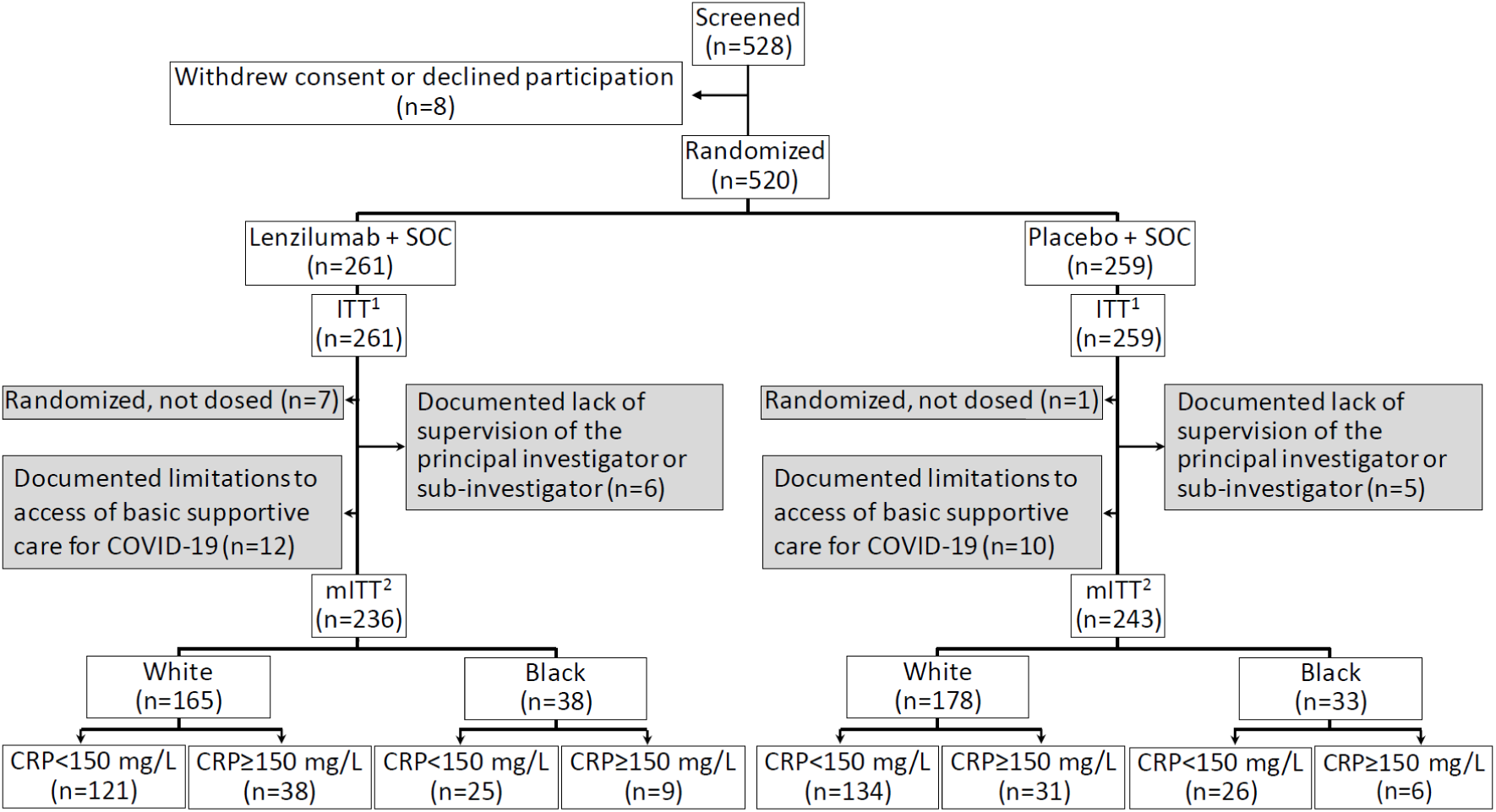
Randomization and Analysis Populations. The ITT population comprised all randomized participants, and the safety set included all participants receiving ≥ 1 dose of study drug. The mITT population comprised the randomized participants who received ≥ 1 dose of study drug under the documented supervision of the principal investigator or sub-investigator and excluded participants from sites that experienced documented limitations to access of basic supportive care for COVID-19.[3] Only patients with a documented CRP level at baseline were included in CRP sub-analyses. Fix the >150 in the chart ^1^ All randomized patients. ^2^ Randomized patients who received at least 1 dose of study drug under the documented supervision of the principal investigator or sub-investigator and excludes site that experienced documented limitations to access of basic supportive care for COVID-19.

The B/AA and White populations were similar in terms of gender and age (Table 1). Approximately 60% of the population was male and the mean age was approximately 60 years. The proportion of patients with a BMI ≥ 30 kg/m^2^ was greater in B/AA participants treated with lenzilumab compared to White participants treated with lenzilumab (Table 1). Greater proportions of B/AA participants self-identified as neither Hispanic nor Latino compared with White participants. A lesser percentage of B/AA participants had more severe COVID-19 at baseline (clinical score=3) than White participants. B/AA participants were generally more at risk of disease progression compared to White participants evidenced by having a higher proportion of comorbidities including hypertension and diabetes. More B/AA participants treated with lenzilumab had congestive heart failure, asthma, and COPD/interstitial lung disease. BB/AA participants, in general, had more risk factors than White participants, particularly in those receiving lenzilumab treatment. The majority of both B/AA and White participants had baseline CRP levels <150 mg/L with similar percentages having CRP≥ 150 mg/L.

**Table 1.** Baseline Characteristics.

|  |  | Black/African American |  | White |  |
| --- | --- | --- | --- | --- | --- |
|  |  | Lenzilumab<br>(n=38) | Placebo<br>(n=33) | Lenzilumab<br>(n=165) | Placebo<br>(n=178) |
| **Gender, n (%)** |  |  |  |  |  |
|  | Female | 15 (39.5) | 14 (42.4) | 56 (33.9) | 59 (33.1) |
|  | Male | 23 (60.5) | 19 (57.6) | 109 (66.1) | 119 (66.9) |
| **Age, years** |  |  |  |  |  |
|  | Mean (SD) | 59.2 (9.7) | 57.5 (12.0) | 61.5 (14.3) | 60.9 (14.35) |
| **BMI** |  |  |  |  |  |
|  | Mean (SD), kg/m<sup>2</sup> | 35.9 (11.2) | 31.3 (7.3) | 32.2 (6.9) | 32.1 (8.0) |
|  | ≥30 kg/m<sup>2</sup>, n (%) | 29 (76.3) | 13 (39.4) | 88 (53.3) | 100 (56.2) |
| **Ethnicity (%)** |  |  |  |  |  |
|  | Hispanic or Latino | 4 (10.5)<sup>a</sup> | 3 (9.1)<sup>a</sup> | 62 (37.6) | 79 (44.4) |
|  | Not Hispanic or Latino | 33 (86.8)<sup>a</sup> | 29 (87.9)<sup>a</sup> | 103 (62.4) | 98 (55.1) |
| **Baseline severity, n (%)** |  |  |  |  |  |
|  | Room air<br>(clinical ordinal score=5) | 1 (3) | 0 (0) | 10 (6.1) | 10 (5.6) |
|  | Low-flow oxygen<br>(clinical ordinal score=4) | 24 (63) | 21 (64) | 85 (51.5) | 92 (51.7) |
|  | High-flow oxygen or<br>NPPV<br>(clinical ordinal score=3) | 13 (34) | 12 (36) | 70 (42.4) | 76 (42.7) |
| **CRP, n (%)** |  |  |  |  |  |
|  | ≥150 mg/L | 9 (23.7) | 6 (18.2) | 38 (23.0) | 31 (17.4) |
|  | <150 mg/L | 25 (65.8) | 26 (78.8) | 121 (73.0) | 134 (75.3) |
| **Comorbidity, n (%)** |  |  |  |  |  |
|  | Hypertension | 30 (78.9) | 26 (78.8) | 98 (59.4) | 119 (66.9) |
|  | Congestive heart failure | 9 (23.7) | 4 (12.1) | 20 (12.1) | 17 (9.6) |
|  | Coronary artery disease | 8 (21.1) | 4 (12.1) | 26 (15.8) | 22 (12.4) |
|  | Diabetes (type 1 and 2) | 30 (78.9) | 24 (72.7) | 74 (44.8) | 92 (51.7) |
|  | Chronic kidney disease | 8 (21.1) | 7 (21.2) | 25 (15.2) | 25 (14.0) |
|  | Chronic Liver disease | 0 (0) | 1 (3) | 9 (5.5) | 12 (6.7) |
|  | Asthma | 7 (18.4) | 1 (3.0) | 21 (12.7) | 17 (9.6) |
|  | COPD/Interstitial lung disease | 7 (18.4) | 3 (9.1) | 13 (7.9) | 15 (8.4) |
| **Number of risk factors**<br>**(excluding age), n (%)** |  |  |  |  |  |
|  | ≥1 | 37 (97) | 32 (97) | 130 (78.8) | 148 (83.1) |
|  | ≥2 | 37 (97) | 30 (91) | 113 (68.5) | 133 (74.7) |
|  | ≥3 | 29 (76) | 18 (55) | 77 (46.7) | 94 (52.8) |
|  | ≥4 | 19 (50) | 10 (30) | 45 (27.3) | 52 (29.2) |

<sup>a</sup> Missing data

### Primary Outcome, Subgroup Analysis

Lenzilumab treatment was associated with a numerically greater likelihood of achieving SWOV (primary endpoint) but did not achieve statistical significance compared to placebo in both B/AA (86.8% vs 70.9%; HR, 2.68; 95% confidence interval [CI], 0.88-8.11; p=0.08; Figure 2A; Table 2) and White participants (85.1% vs 80.8%; HR, 1.41; 95%CI, 0.85-2.35, p=0.18; Figure 2B; Table 2). When baseline CRP<150 mg/mL was considered, SWOV was significantly improved in both B/AA participants (HR: 8.9; 95%CI: 1.08, 73.09; p=0.04; Figure 3A; Table 2) and White participants (HR: 2.32; 95%CI: 1.17, 4.61; p=0.02; Figure 3B; Table 2). The number needed to treat was calculated to be 4 in B/AA participants and 12 in White participants in whom baseline CRP was <150 mg/L (Table 2). SWOV was not improved in participants of either race with baseline CRP150≥ mg/L (Table 2; Figure 3C and D).

**Table 2.** Survival Without Ventilation (SWOV), Primary Endpoint^a,b^.

|  | Black/African American |  | White |  |
| --- | --- | --- | --- | --- |
|  | Lenzilumab | Placebo | Lenzilumab | Placebo |
| **Overall** |  |  |  |  |
| % (95% CI) | 86.8<sup>c</sup> (71.2, 94.3)<br>(n=38) | 70.9 (51.5, 83.7)<br>(n=33) | 85.1 (78.5, 89.7)<br>(n=165) | 80.8 (74.2, 85.9)<br>(n=178) |
| HR (95% CI)<sup>d</sup> | 2.68 (0.88, 8.11) |  | 1.41 (0.85, 2.35) |  |
| p value | 0.08 |  | 0.18 |  |
| **CRP<150 mg/L** |  |  |  |  |
| % (95% CI) | 96.0 (74.8, 99.4)<br>(n=25) | 70.7 (48.1, 84.9)<br>(n=26) | 90.6 (83.7, 94.7)<br>(n=121) | 82.0 (74.3, 87.5)<br>(n=134) |
| HR (95% CI)<sup>d</sup> | 8.90 (1.08, 73.09) |  | 2.32 (1.17, 4.61) |  |
| p value | 0.04 |  | 0.02 |  |
| Number Needed to Treat | 4 |  | 12 |  |
| **CRP≥150 mg/L** |  |  |  |  |
| % (95% CI) | 77.8 (37.0, 93.9)<br>(n=9) | 83.3 (27.3, 97.5)<br>(n=6) | 70.4 (52.9, 82.4)<br>(n=38) | 71.0 (51.7, 83.7)<br>(n=31) |
| HR (95% CI)<sup>d</sup> | 0.63 (0.06, 6.91) |  | 1.034 (0.44, 2.44) |  |
| p value | 0.70 |  | 0.94 |  |

<sup>a</sup> All data censored at 28 days following enrollment.
<sup>b</sup> mITT, modified intention to treat population.
<sup>c</sup> Kaplan-Meier estimates for proportion of participants.
<sup>d</sup> Cox proportional hazard model for time to event with age ( $\leq 65$ , $>65$ ) and severity (severe, critical strata) as covariates.

**Figure 2.**
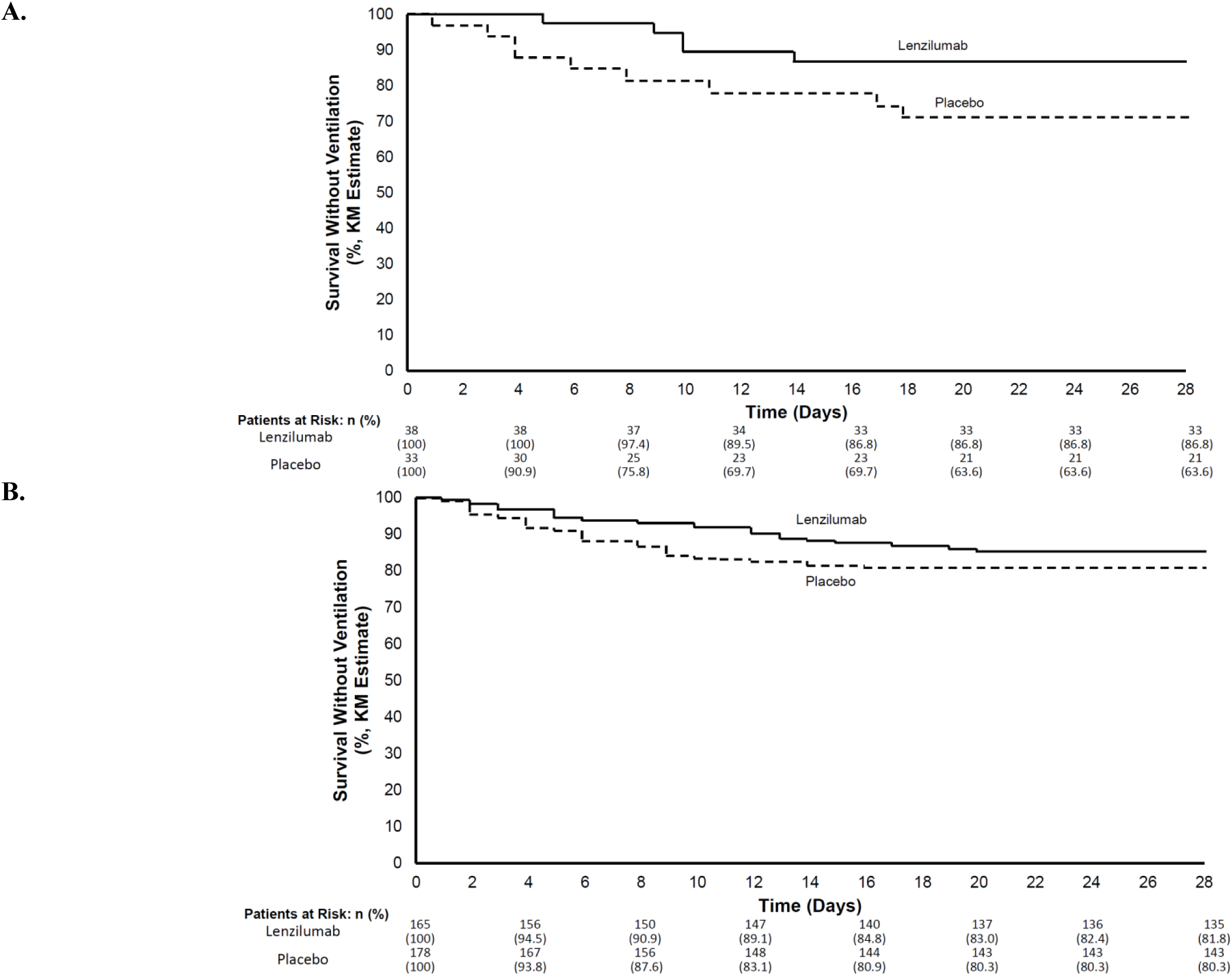
Survival Without Ventilation Through Day 28 (mITT). Kaplan-Meier curves representing: **A**. B/AA participants (n=71). B. White participants (n=343)

**Figure 3.**
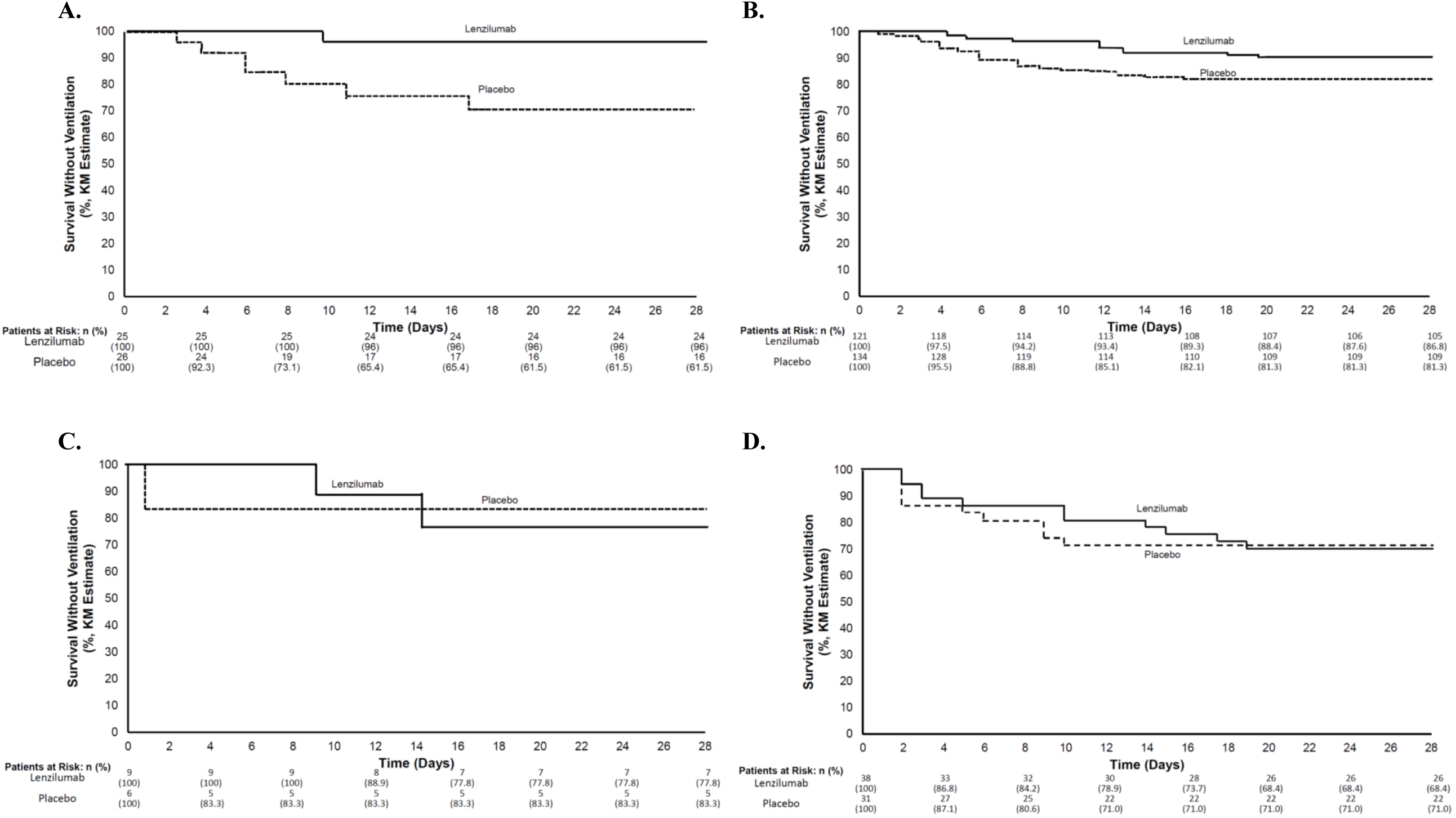
Survival Without Ventilation Through Day 28 According to Baseline CRP (mITT). Kaplan-Meier curves representing: **A**. B/AA participants with baseline CRP < 150 mg/L (n=51). **B**. White participants with baseline CRP < 150 mg/L (n=155). **C**. B/AA participants with baseline CRP ≥ 150 mg/L (n=15). **D**. White participants with baseline CRP ≥ 150 mg/L (n=69).

### Key Secondary Endpoints, Subgroup Analysis

The incidence of IMV, ECMO or mortality; ventilator-free days; ICU days and time to recovery were not significantly different between lenzilumab-or placebo-treated patients in participants of either race, although the endpoints were numerically improved with lenzilumab treatment (Table 3). In B/AA participants, IMV, ECMO or mortality occurred in 13.9% of those treated with lenzilumab compared to 29.3% of those treated with placebo (OR, 0.39; 95% CI. 0.11-1.34; *P*=0.14) and in White participants, IMV, ECMO, or mortality occurred in 13.3% of patients treated with lenzilumab compared to 17.7% of those treated with placebo (OR, 0.71; 95% CI. 0.39-1.32; *P*=0.28). Mortality was numerically less in lenzilumab-treated B/AA participants (7.9%) compared with placebo (16.5%) but not in lenzilumab-treated White participants (10.6%) compared with their respective placebo-treated (11.4%) group. Both B/AA and White participants treated with lenzilumab had numerically fewer mean ICU days than those who received placebo, though this was not statistically significant. Ventilator-free days were not different between lenzilumab and placebo treatment in either race (Table 3).

**Table 3.**
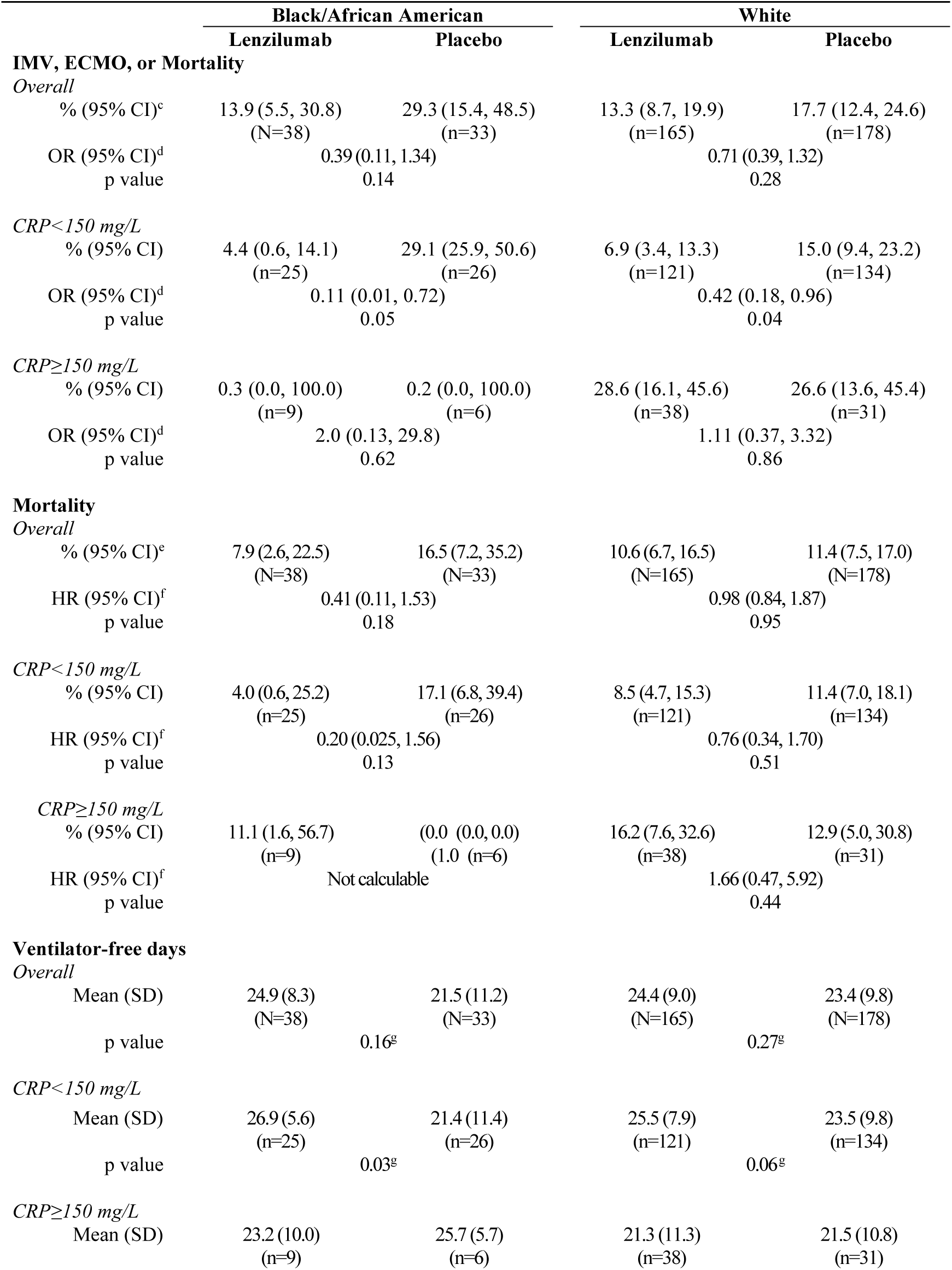
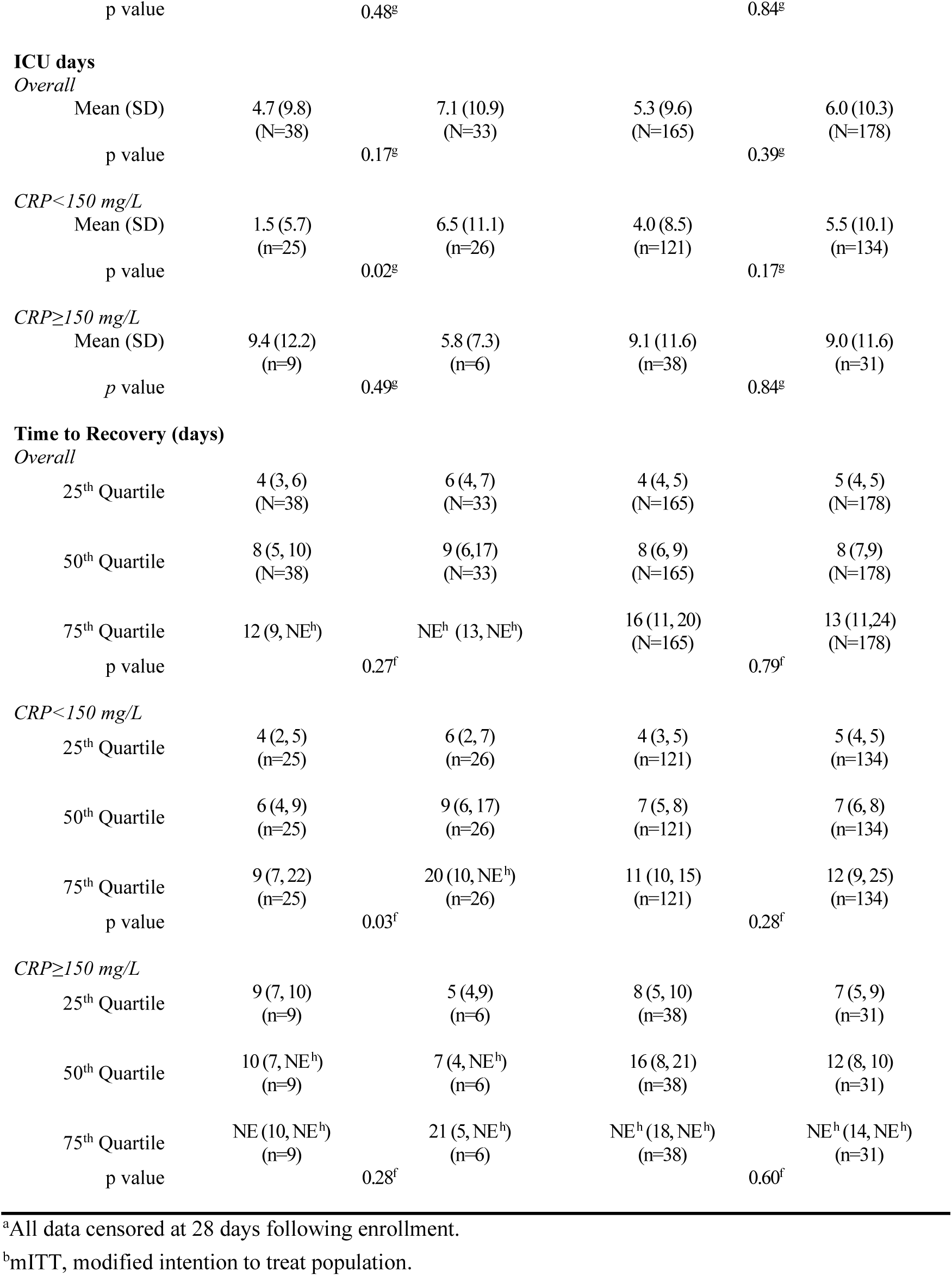
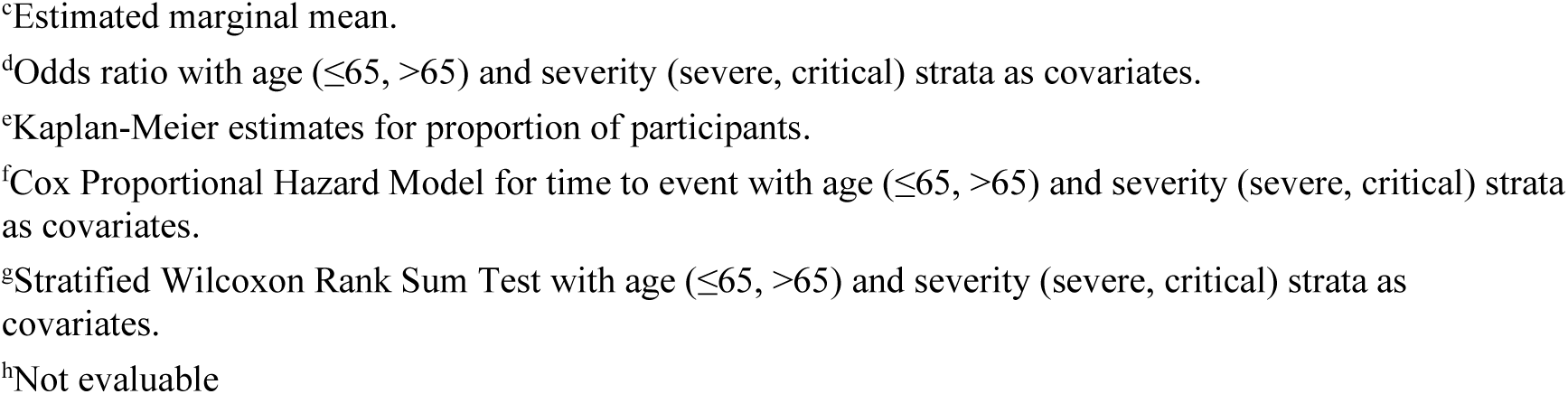
Key Secondary Endpoints^a,b^.

Participants of both races with CRP<150 mg/L had a greater improvement in key secondary endpoints than observed in the overall population (Table 2). The incidence of IMV, ECMO or mortality was less (4.4%) in B/AA participants with CRP<150 mg/L and treated with lenzilumab compared with those treated with placebo (29.1%; OR, 0.11; 0.01,0.72; p=0.05). IMV, ECMO or mortality occurred in 6.9% of lenzilumab-treated and 15.0% placebo-treated White participants (OR, 0.42; 0.18, 0.96, p=0.04). Similar numerical improvements were observed in mortality (B/AA participants, p=0.13; White participants p=0.51); ventilator free days (B/AA participants, p=0.03; White participants p=0.51); ICU days (B/AA participants, p=0.02; White participants p=0.17); and time to recovery (B/AA participants, p=0.03; White participants p=0.28).

### Safety

In the safety population, adverse events ≥ Grade 3 were reported in 23.8% to 32.4% of the participants treated with lenzilumab or placebo (Table 4). Respiratory, thoracic, and mediastinal disorders were the most common adverse event followed by cardiac disorders. In White participants no differences in adverse events were observed between lenzilumab- and placebo-treated participants. Small sample size in the B/AA participants group precluded meaningful statistical comparison of adverse events, but generally no differences were apparent. No infusion-related reactions or serious adverse events; including, hematologic laboratory abnormalities, liver enzyme abnormalities, increased incidence of infection, or cases of pulmonary alveolar proteinosis were reported with lenzilumab treatment.

**Table 4.** Most Common Grade ≥ 3 Adverse Events (Overall Incidence ≥ 1.0%)

| **System Organ Class**<br>**Preferred term n (%)** | **Black / African American** |  | **White** |  |
| --- | --- | --- | --- | --- |
|  | **Lenzilumab**<br>**(N=42)** | **Placebo**<br>**(n=37)** | **Lenzilumab**<br>**(n=177)** | **Placebo**<br>**(n=185)** |
| Any AE ≥ Grade 3 | 10 (23.8) | 12 (32.4) | 39 (22.0) | 46 (24.9) |
| Respiratory, thoracic, and mediastinal disorders | 9 (21.4) | 10 (27.0) | 37 (20.9) | 43 (23.2) |
| Respiratory failure | 5 (11.9) | 5 (13.5) | 15 (8.5) | 21 (11.4) |
| Acute respiratory failure | 2 (4.8) | 4 (10.8) | 12 (6.8) | 14 (7.6) |
| Pulmonary embolism | 2 (4.8) | 1 (2.7) | 1 (1.1) | 1 (0.5) |
| Hypoxia | 0 (0.0) | 1 (2.7) | 7 (4.0) | 6 (3.2) |
| Acute respiratory distress syndrome | 1 (2.4) | 0 (0.0) | 2 (1.1) | 2 (1.1) |
| Cardiac disorders | 4 (9.5) | 2 (5.4) | 7 (4.0) | 10 (5.4) |
| Cardiac arrest | 3 (7.1) | 1 (2.7) | 4 (2.3) | 2 (1.1) |
| Cardio-respiratory arrest | 1 (2.4) | 1 (2.7) | 2 (1.1) | 3 (1.6) |
| Renal and urinary disorders | 0 (0.0) | 3 (8.1) | 2 (1.1) | 4 (2.2) |
| Acute kidney injury | 0 (0.0) | 2 (5.4) | 2 (1.1) | 3 (1.6) |
| Renal failure | 0 (0.0) | 1 (2.7) | 0 (0.0) | 0 (0.0) |
| Vascular disorders | 2 (4.8) | 2 (5.4) | 2 (1.1) | 5 (2.7) |
| Shock | 1 (2.4) | 0 (0.0) | 0 (0.0) | 2 (1.1) |
| Hypotension | 0 (0.0) | 2 (5.4) | 0 (0.0) | 0 (0.0) |
| Peripheral artery occlusion | 1 (2.4) | 0 (0.0) | 0 (0.0) | 0 (0.0) |
| Shock hemorrhagic | 0 (0.0) | 0 (0.0) | 0 (0.0) | 2 (1.1) |
| Infections and infestations | 0 (0.0) | 1 (2.7) | 8 (4.5) | 6 (3.2) |
| Sepsis | 0 (0.0) | 1 (2.7) | 1 (0.6) | 2 (1.1) |
| Pneumonia | 0 (0.0) | 0 (0.0) | 2 (1.1) | 1 (0.5) |
| Septic shock | 0 (0.0) | 0 (0.0) | 4 (2.3) | 5 (2.7) |
| Pneumonia, bacterial | 0 (0.0) | 0 (0.0) | 0 (0.0) | 2 (1.1) |
| Gastrointestinal disorders | 0 (0.0) | 1 (2.7) | 1 (0.6) | 2 (1.1) |
| Diverticular perforation | 0 (0.0) | 1 (2.7) | 0 (0.0) | 0 (0.0) |
| Upper gastrointestinal hemorrhage | 0 (0.0) | 0 (0.0) | 0 (0.0) | 2 (1.1) |
| General disorders and administration site conditions | 1 (2.4) | 1 (2.7) | 2 (1.1) | 3 (1.6) |
| Multiple organ dysfunction syndrome | 1 (2.4) | 1 (2.7) | 1 (0.6) | 2 (1.1) |

## DISCUSSION

Epidemiologic studies demonstrate that in the United States and in Brazil, B/AA patients have a higher risk of contracting the SARS-CoV-2 virus and dying from COVID-19.[5-7] The current sub-analysis of LIVE-AIR sought to characterize the efficacy of lenzilumab in both the B/AA and White subpopulations, particularly when treated in the earlier stages of COVID-19 progression. LIVE-AIR included a proportion of B/AA participants that was slightly greater percentage than observed in the general US population. SWOV was numerically improved to a greater extent in lenzilumab-treated B/AA participants than in White participants, when each were compared with their matched controls, respectively. SWOV was significantly improved in participants with CRP<150 mg/L with a much greater effect in B/AA participants. The results reinforce the importance of early treatment in the hyperinflammatory immune response and/or maintaining a slower/milder trajectory of COVID-19 progression, guided by plasma CRP as previously reported from LIVE-AIR.[4]

Social determinants of health augment the burden of COVID-19-related illness in B/AA as evidenced by more infections and hospitalizations, and higher mortality rates than Whites.[5-7] Indeed in this analysis, B/AA exhibited greater proportion of participants with co-morbid hypertension, diabetes, chronic kidney disease, two or more co-morbid conditions, and with some co-morbidities a greater burden in the lenzilumab treatment group. Despite the greater health burden in B/AA, lenzilumab resulted in greater improvement in SWOV in this group than in Whites, especially when baseline CRP<150 mg/L. By extrapolation, lenzilumab appeared to decrease the hyperinflammatory immune response in B/AA, in whom exists a greater risk of adverse outcomes due to a multiplicity of baseline factors including or resulting from social determinants of health, thus creating health equity. Indeed, worse outcomes were observed in B/AA compared with Whites in the absence of lenzilumab treatment (Tables 2 and 3).

Given that LIVE-AIR was a well-controlled interventional study in which the standard of care was rigorously maintained across the study and any participant not so managed was excluded from the mITT analysis, biologic differences either resulting from or contributing to social determinants of health possibly contribute to differences in lenzilumab and placebo treatment between B/AA and White participants. Although any reason is speculative, several may be candidates. The greater prevalence of ACE deletions in B/AA is postulated to contribute to poor health outcomes in part through chronic inflammatory processes proposed to be augmented by SAR-CoV-2 infection through the ACE2 receptor[11], although this has yet to be demonstrated empirically in COVID-19. G6PD deficiency, which is more prevalent in B/AA, permits propagation of tissue damage through reactive oxygen species that are generated with hyperinflammation.[19] Both pathways involve GM-CSF signaling as demonstrated by preclinical investigations and analyses of the associated clinical variables.[20-25] The Duffy null phenotype estimated to be present in approximately 67% of African Americans[14] may result in the failure to sequester[15] an abundance of proinflammatory cytokines including GM-CSF triggered by SARS-CoV-2 infection thereby propagating hyperinflammatory immune response. While evidence supports the putative role of inflammation in these pathways, other pathways may be elucidated by which B/AA are more susceptible and affected by COVID-19.

Several limitations exist with the analyses performed herein. As with any subgroup analysis, there may be statistical challenges, including multiple comparisons and inadequate power. No statistical comparisons were made between B/AA and White subpopulations, due to small sample sizes which limits the interpretability of the data presented. Additionally, the sub-analysis was *post-hoc* and not pre-specified. Given that the study was conducted in the United States and Brazil, inter- and intra-country differences in standard of care may vary, including but not limited to access to remdesivir. Furthermore, many cases in Brazil were enrolled during the surge when there were documented limitations to access basic supportive COVID-19 care, including high-flow oxygen devices; this resulted in a disproportionate increase from low-flow supplemental oxygen directly to IMV. Because of these limitations the findings herein require independent confirmation. Regardless of the limitations, the findings suggest an improvement in equity of outcomes with lenzilumab treatment.

The COVID-19 pandemic exhibits disparities in outcomes related to race. An understanding of the social and biologic determinants that intersect to affect COVID-19 transmission and outcomes is evolving. The subgroup analysis of B/AA and White subpopulations from the LIVE-AIR study affirms results observed in the general population that blocking GM-CSF activity with lenzilumab improved outcomes in adults with COVID-19, especially in those with baseline CRP<150 mg/L. The results also suggest an unexpectedly greater improvement in outcomes in the B/AA subpopulation, a hypothesis for which could be related to biologic mechanisms and driven by SDOH. Additional prospective investigation is required to further expand on these results.

## Data Availability

All data produced in the present work are contained in the manuscript

## ACKNOWLEDGMENTS

The authors thank all participants and investigators in LIVE-AIR and RxMedical Dynamics, LLC for its editorial support.

